# Assessing Brain Involvement in Fabry Disease with Deep Learning and the Brain-Age Paradigm

**DOI:** 10.1101/2023.04.04.23288000

**Authors:** Alfredo Montella, Mario Tranfa, Alessandra Scaravilli, Frederik Barkhof, Arturo Brunetti, James Cole, Michela Gravina, Stefano Marrone, Daniele Riccio, Eleonora Riccio, Carlo Sansone, Letizia Spinelli, Maria Petracca, Antonio Pisani, Sirio Cocozza, Giuseppe Pontillo

## Abstract

**Background:** While neurological manifestations are core features of Fabry disease (FD), quantitative neuroimaging biomarkers allowing to measure brain involvement are lacking. We used deep learning and the brain-age paradigm to assess whether FD patients’ brains appear older than normal and to validate brain-predicted age difference (brain-PAD) as a possible disease severity biomarker.

**Methods:** MRI scans of FD patients and healthy controls (HC) from a single Institution were retrospectively studied. The Fabry stabilization index (FASTEX) was recorded as a measure of disease severity. Using minimally preprocessed 3D T1-weighted brain scans of healthy subjects from 8 publicly available sources (N=2160; mean age=33y[range 4-86]), we trained a model predicting chronological age based on a DenseNet architecture and used it to generate brain-age predictions in the internal cohort. Within a linear modeling framework, brain-PAD was tested for age/sex-adjusted associations with diagnostic group (FD vs HC), FASTEX score, and both global and voxel-level neuroimaging measures.

**Results:** We studied 52 FD patients (40.6±12.6y; 28F) and 58 HC (38.4±13.4y; 28F). The brain-age model achieved accurate out-of-sample performance (mean absolute error=4.01y, R^2^=0.90). FD patients had significantly higher brain-PAD than HC (estimated marginal means: 3.1vs-0.1, p=0.01). Brain-PAD was associated with FASTEX score (B=0.10, p=0.02), brain parenchymal fraction (B=-153.50, p=0.001), white matter hyperintensities load (B=0.85, p=0.01), and tissue volume reduction throughout the brain.

**Conclusions:** We demonstrated that FD patients’ brains appear older than normal. Brain-PAD correlates with FD-related multi-organ damage and is influenced by both global brain volume and white matter hyperintensities, offering a comprehensive biomarker of (neurological) disease severity.

**Summary Statement:** Using deep learning and the brain-age paradigm, we found that Fabry disease is associated with older-appearing brains. The gap between brain-predicted and chronological age correlates with multi-organ disease severity, offering a novel quantitative neuroimaging biomarker.

**Key Points:**

- Patients with Fabry disease show significantly higher brain-predicted age difference values compared to healthy controls (estimated marginal means: 3.1 vs -0.1, p=0.01).
- Brain-predicted age difference correlates with multi-organ disease severity and is associated with brain parenchymal fraction, white matter hyperintensities load, and tissue volume throughout the brain.
- Brain-predicted age difference might represent a sensitive quantitative biomarker of brain involvement in Fabry disease, with potentially relevant implications for patient stratification and treatment response monitoring.

## Introduction

Fabry disease (FD; OMIM 301500) is a rare X-inherited lysosomal storage disorder characterized by the accumulation of metabolites in various cell types, resulting from the absent or markedly deficient activity of the enzyme α-galactosidase A (α-Gal A) and leading to damage and loss of function of especially the kidney, heart, and brain.(1)

Involvement of the central nervous system is mainly characterized by vascular pathology, the severity of which may greatly vary, reflecting the complex pathophysiology of tissue damage in FD.(2) However, while the recommended follow-up of patients with FD includes brain MR imaging, an accurate evaluation of FD-related brain damage is hampered by the lack of quantitative imaging biomarkers,(3) which also limits the possibility to monitor the effectiveness of recently introduced specific treatments on cerebral manifestations.(4)

In the search for objective imaging-derived markers of brain health and pathology, the brain-age paradigm has emerged as a promising approach.(5) Briefly, machine learning methods are used to model chronological age as a function of structural brain MRI scans in healthy people, and the resulting model of ‘normal’ brain aging is used for neuroimaging-based age prediction in unseen subjects.(5) The extent to which each subject deviates from healthy brain-aging trajectories, expressed as the difference between predicted and chronological age (the brain-predicted age difference, brain-PAD), has been proposed as an index of structural brain health, sensitive to pathology in a wide spectrum of neurological and psychiatric disorders.(6) As a relevant example, brain-age predictions are sensitive to white matter hyperintensities (WMH) and brain volumes, imaging features that are both manifestations of cerebral small vessel disease,(7–9) which is thought to be the main pathogenetic mechanisms through which FD impacts brain health.(2)

Here, we applied the brain-age paradigm to investigate brain involvement in patients with FD. Our main aims were: i) to assess whether FD brains deviate from normal aging trajectories; ii) to validate brain-PAD as a measure of disease severity against other established clinical markers; iii) to explore the neuroimaging determinants of brain-age prediction in this condition.

## Materials and Methods

### Participants

In this retrospective, cross-sectional study, part of a larger monocentric research program on FD,(10–13) patients with a genetically confirmed diagnosis were selected,(14) along with age- and sex-comparable healthy controls (HC). Participants with history of major cerebrovascular events were excluded. Additional exclusion criteria were age <18 or >65 years, and the presence of other relevant neurological or systemic conditions.

Scores quantifying the involvement of nervous, renal and cardiac systems in FD patients were computed based on clinical variables recorded within 1 month from the MRI and summed to obtain a cumulative measure of multi-organ damage severity, the total raw Fabry stabilization index (FASTEX) score, ranging from 0 (normal) to 28 (maximum severity).(15)

The study was conducted in compliance with ethical standards and approved by the local ethics committee “Carlo Romano”. Written informed consent was obtained from all participants according to the Declaration of Helsinki.

### MRI acquisition and preprocessing

All MRI examinations were acquired between October 2015 and April 2019 using the same 3T scanner (Magnetom Trio, Siemens Healthineers), equipped with an 8-channel head coil. The acquisition protocol included a 3D T1-weighted (T1w) magnetization prepared rapid acquisition gradient echo (MPRAGE) sequence (TR=1900 ms; TE=3.4 ms; TI=900 ms; flip angle=9°; voxel size=1×1×1 mm^3^; 160 axial slices) and, for FD patients, a 3D T2-weighted fluid attenuated inversion recovery (FLAIR) sequence for WMH assessment (TR=6000 ms; TE=396 ms; TI=2200 ms; Flip Angle=120°; voxel size=1×1×1 mm^3^; 160 sagittal slices).

For FD patients, WMH were automatically segmented on FLAIR images using Lesion Segmentation Tool v3.0.0 (www.statistical-modelling.de/lst.html) and used to fill lesions in T1w images. For all participants, we used the Computational Anatomy Toolbox v12.8 (http://www.neuro.uni-jena.de/cat) to segment T1w volumes into grey matter (GM), white matter (WM) and cerebrospinal fluid (CSF) and obtain MNI-normalized, modulated, smoothed (1mm full-width at half-maximum isotropic Gaussian kernel) GM and WM probability maps Summary measures of GM, WM, CSF, and total intracranial (TIV) volumes were also generated, and brain parenchymal fraction (BPF) was computed as the ratio of brain volume to TIV.

### Brain-age modelling

A model of healthy brain aging was trained and evaluated on a large external dataset (total N=2160; male/female=1293/867; mean age=33 years, age range=4-86) comprising 3D T1-weighted brain scans of healthy subjects from 8 publicly available sources (Supplementary Table 1).

Raw T1w volumes underwent minimal preprocessing, including DICOM to NIfTI conversion, correction for intensity non-uniformity with N4 bias field correction,(16) rigid registration to the MNI space and resampling to 1.5 mm^3^ voxels, to ensure consistency of spatial orientation and resolution. Finally, images were intensity-normalized by subtracting the image mean and dividing by the image standard deviation.

Our brain-age model was based on the DenseNet264 architecture,(17) adapted from the implementation available at Project MONAI (https://docs.monai.io/en/stable/_modules/monai/networks/nets/densenet.html) by adding a linear regression layer for the prediction of a continuous variable and a 0.2 dropout rate after each dense layer to reduce the risk of overfitting. Modeling was performed with PyTorch 1.12.0(18) using one NVIDIA Tesla V100S 32 GB graphics processing unit. The full dataset was randomly split into training (64%=1382), validation (16%=346) and test (20%=432) sets (Supplementary Figure 1). Mean absolute error (MAE) and coefficient of determination (R^2^) were used to quantify model performance. Lastly, age bias (i.e., underestimation of age in older subjects and vice versa) was statistically corrected as in *de Lange et al*.,(19) and the final model was applied to the internal cohort of FD patients and HC to generate brain-predicted ages and corresponding brain-PAD values.

An outline of the different steps of the brain-age modelling procedure is displayed in Figure 1.

**Figure 1.**
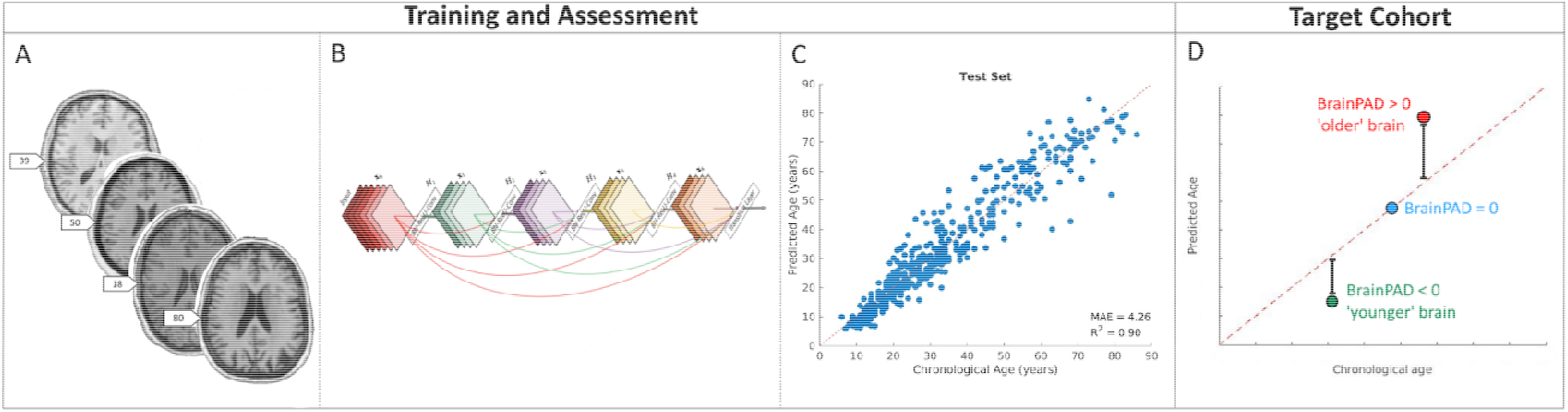
Outline of the brain-age modeling procedure. Minimally preprocessed T1-weighted images (A) are used as input for the training and the evaluation of a model for the prediction of chronological age based on a 3D DenseNet architecture (B). The model with the lowest validation loss is chosen, and performance is measured on the previously unseen cases of the test set (C). The final model is also applied to the target clinical population (D), composed of the internal cohort of FD patients and HC, to generate brain-predicted ages and corresponding brain-PAD values. FD: Fabry Disease; HC: Healthy Controls; brain-PAD: brain-predicted age difference.

### Statistical analysis

Unless otherwise specified, statistical analyses were carried out using the Statistical Package for Social Science (SPSSv25.0, IBM corp.), with a statistical significance level α=0.05 and 95% confidence intervals (CI) and *p* values computed using bootstrap with 1000 resamples.

To assess possible between-group differences in terms of brain-PAD, we used one-way ANCOVA, controlling for the effects of age, age^2^ and sex and calculating estimated marginal means for the two groups.

To validate brain-PAD as a measure of disease severity, we tested its association with the FASTEX score in a linear regression model including also age, age^2^ and sex.

To investigate the neuroimaging determinants of brain-PAD in patients with FD, we used hierarchical linear regression analyses with age, age^2^ and sex in the first block and WMH load or BPF in the second block. Similarly, we tested age-, age^2^- and sex-adjusted associations between brain-PAD and TIV-scaled, preprocessed GM and WM maps, using a nonparametric approach based on 5000 permutations applied to the general linear model(20) via the Threshold Free Cluster Enhancement toolbox (http://www.neuro.uni-jena.de/tfce). The same analysis was repeated after adding the variables group and group*brain-PAD interaction in the model, with this latter term intended to test the hypothesis that different voxel-wise patterns might influence brain-age prediction in the two groups.

## Results

A flow diagram summarizing the patient selection procedure is shown in Figure 2. A total of 52 patients with FD were identified (40.6 ± 12.6 years; M/F: 24/28), along with 58 HC (38.4 ± 13.4 years; M/F: 30/28). The median FASTEX score of FD patients was 6 (range 3 – 9), indicating mild/to/moderate involvement. Complete demographic, clinical, and MRI characteristics of the studied population are available in Table 1.

**Figure 2.**
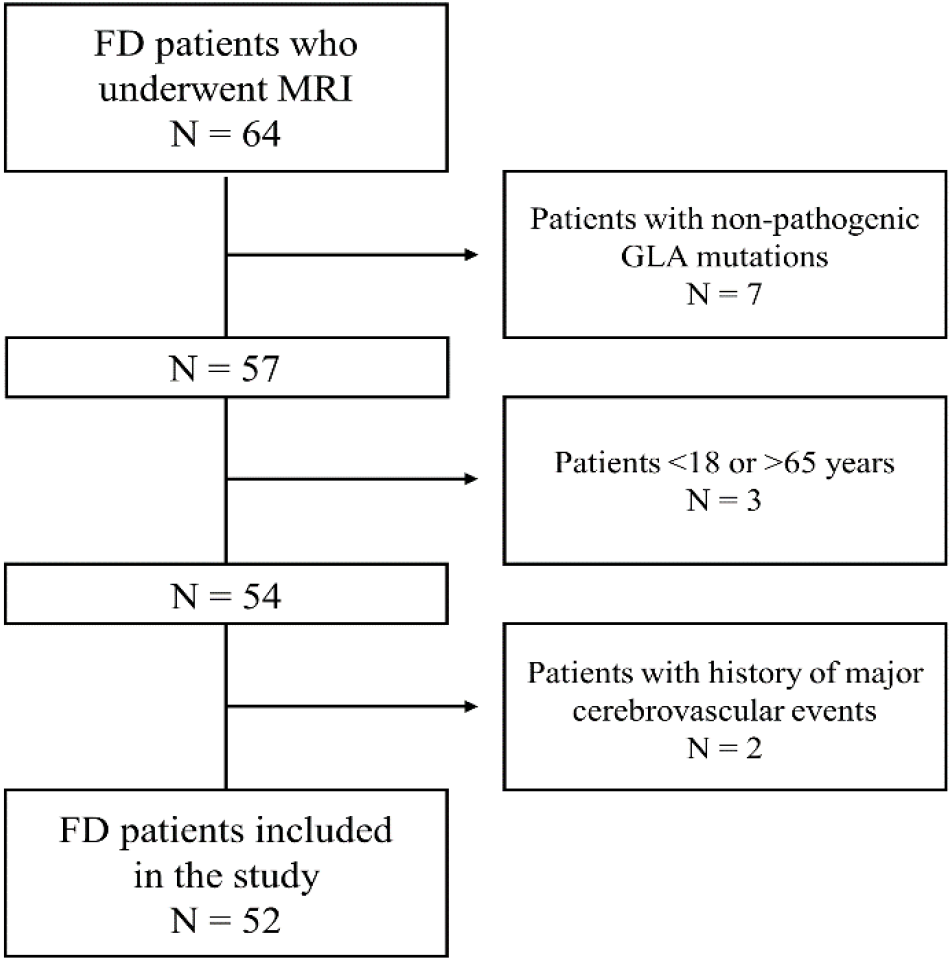
Flow diagram summarizing the patient selection procedure.

**Table 1.** Demographic, clinical, and MRI characteristics of all the participants included in the study.

|  | FD<br>N = 52 | HC<br>N = 58 | *p* value<br>(FD vs HC) |
| --- | --- | --- | --- |
| Age (y) | 40.6 ± 12.6 | 38.4 ± 13.4 | 0.37 |
| Sex (M/F)* | 24/28 | 30/28 | 0.56 |
| FASTEX score† |  |  |  |
| Total score | 6 (3 – 9) | n.a. | n.a. |
| Nervous system score | 2 (1 – 2) | n.a. | n.a. |
| Renal system score | 1 (1 – 3) | n.a. | n.a. |
| Cardiac system score | 2 (1 – 3) | n.a. | n.a. |
| BPF | 0.81 ± 0.03 | 0.82 ± 0.03 | 0.06 |
| WMH load (ml) | 0.9 ± 3.7 | n.a. | n.a. |

Unless otherwise specified, data are expressed as mean ± standard deviation. Between-group differences were tested with either Student *t* (age and BPF) or chi-square (sex) tests.
\*Expressed as number of subjects.
†Expressed as median, with interquartile range in parentheses.
FD: Fabry Disease; HC: Healthy Control; SD: Standard Deviation; BPF: Brain Parenchymal Fraction; WMH: White Matter Hyperintensities; n.a.: not applicable.

The brain-age model trained on healthy subjects’ MRI scans achieved an accurate out-of-sample age prediction (test set MAE = 4.01 years, R^2^ = 0.90).

When looking at brain-PAD values in the internal cohort, there was a significant effect of diagnostic group when controlling for age, age^2^, and sex (F[1, 105] = 6.46, *p* = 0.01, partial η^2^ = 0.06), with FD patients showing higher values than HC (estimated marginal means 3.1 [95% CI = 1.0 – 5.3] vs -0.1 [95% CI = -1.9 – 1.4]) (Figure 3).

**Figure 3.**
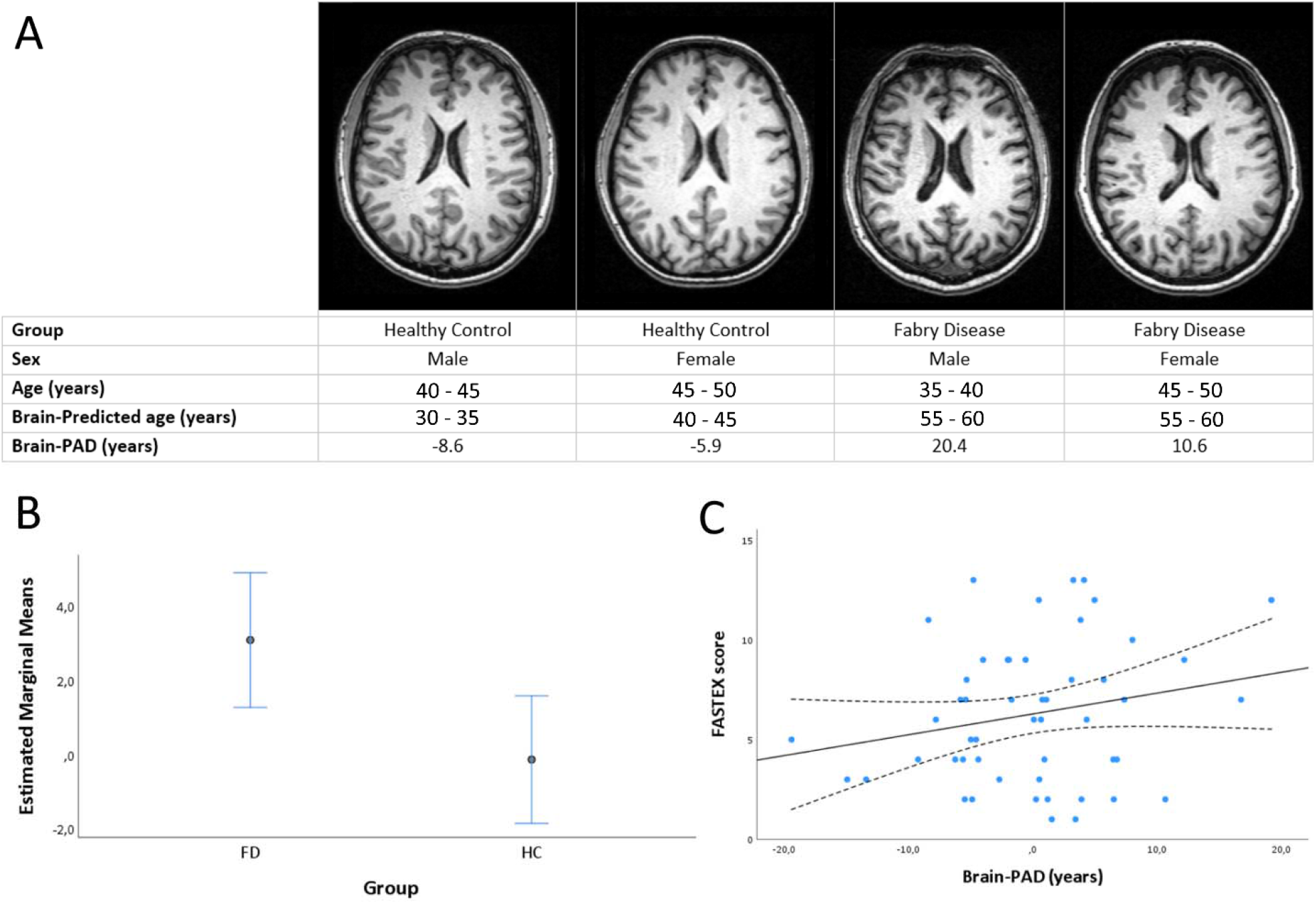
Brain-age prediction in the internal cohort and its relationship with disease status (FD vs HC) and the FASTEX score. In (A), presented are comparable-level axial slices from four example subjects (two per group) with extreme brain-PAD values. In (B), age, age^2^, and sex-adjusted estimated marginal means for the two groups, along with 95% bootstrap confidence intervals. In (C), scatterplot showing the relationship between brain-PAD values (age, age^2^, and sex-adjusted) and FASTEX score in patients with FD. FD: Fabry disease; HC: Healthy Controls; FASTEX: Fabry stabilization index; brain-PAD: brain-predicted age difference.

Brain-PAD was significantly associated with the FASTEX score (B = 0.10 [95% CI = 0.02 – 0.19]; standard error B = 0.04; *p* = 0.02), in a linear model including also age, age^2^ and sex (R^2^ = 0.41, *p* < 0.001) (Figure 3), corresponding to a 0.10 increase in the FASTEX score for each additional year of brain-predicted age.

As for the neuroimaging determinants of brain-PAD in FD patients, both higher WMH load (*p* = 0.01) and lower BPF (*p* = 0.001) were associated with older-appearing brains (Table 2). Voxel-wise, we found a significant inverse correlation between brain-PAD values and tissue volumes diffusely throughout the brain, with the strongest effect sizes observed at the level of the deep and periventricular WM (Figure 4). The interaction analysis revealed no significant effect of diagnostic group on the relationship between local tissue volumes and brain-PAD.

**Table 2.** Results of the hierarchical linear regression analyses for the prediction of brain-PAD in FD patients. Confidence intervals, standard errors, and *p* values are based on 1000 bootstrap samples.

|  | **B (95% CI)** | **SE B** | ***p*** |
| --- | --- | --- | --- |
| **Model 1** |  |  |  |
| Constant | 10.48 (-6.00 – 35.60) | 9.41 | 0.23 |
| Age | -0.12 (-1.19 – 0.71) | 0.54 | 0.82 |
| Age<sup>2</sup> | -0.002 (-0.016 – 0.017) | 0.007 | 0.75 |
| Sex | 1.65 (-2.48 – 6.14) | 2.13 | 0.45 |
| **Model 2** |  |  |  |
| Constant | 8.53 (-7.32 – 39.21) | 8.85 | 0.29 |
| Age | 0.04 (-0.94 – 0.73) | 0.50 | 0.93 |
| Age<sup>2</sup> | -0.005 (-0.017 – 0.012) | 0.006 | 0.43 |
| Sex | 0.53 (-3.37 – 4.13) | 2.04 | 0.80 |
| WMH load | 0.85 (-2.35 – 6.78) | 1.72 | 0.01 |
| **Model 3** |  |  |  |
| Constant | 148.47 (77.24 – 224.90) | 36.81 | 0.001 |
| Age | -0.54 (-1.76 – 0.27) | 0.54 | 0.30 |
| Age<sup>2</sup> | 0.000 (-0.012 – 0.017) | 0.007 | 0.96 |
| Sex | 0.71 (-3.21 – 4.86) | 1.91 | 0.73 |
| BPF | -153.50 (-236.42 – -73.24) | 38.74 | 0.001 |

$R^2$ is 0.18 for Model 1, 0.32 ( $\Delta R^2 = 0.14$ ) for Model 2, and 0.43 ( $\Delta R^2 = 0.25$ ) for Model 3.
CI = Confidence Interval; SE = Standard Error; WMH = White Matter Hyperintensities; BPF = Brain Parenchymal Fraction.

**Figure 4.**
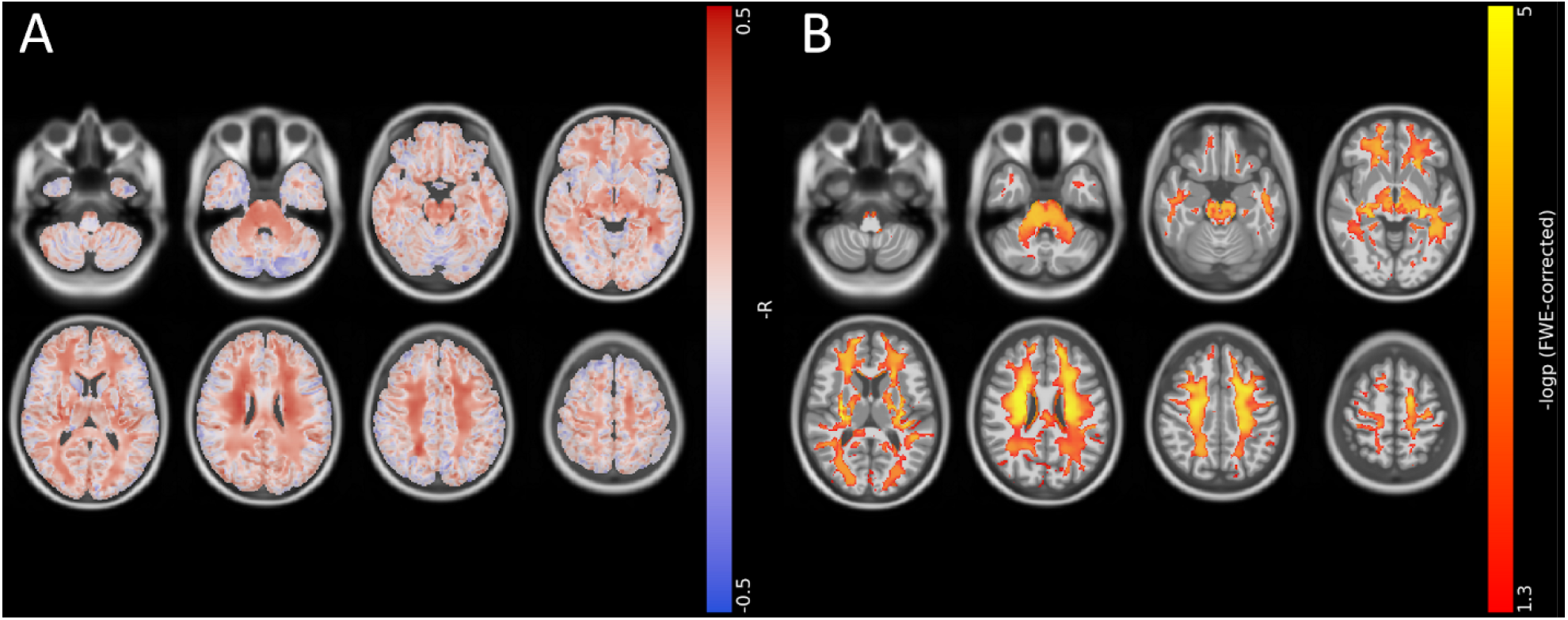
Voxel-wise correlation between brain tissue volumes and brain-PAD in patients with FD. (A) Effect size (-R, *red* to *blue*) and (B) thresholded statistical (-logp, *yellow* to *red*) maps are shown, superimposed on axial sections of a 3D T1-weighted template in standard space. FD: Fabry Disease; brain-PAD: brain-predicted age difference.

## Discussion

By applying the brain-age paradigm in a relatively large cohort of patients, we found that FD is associated with older-appearing brains (on average, 3 years more than normal), indicative of accelerated cerebral aging. The brain-PAD metric correlates with FD-related multi-organ damage and is influenced by both global brain volume and WMH load.

Our brain-age model was built using an ‘off-the-shelf’ standard DenseNet264 configuration, to ensure reproducibility and ease of use. Interestingly, not only the achieved performance was close to that of literature benchmarks (21,22), but the model of healthy brain aging was also sensitive to FD-related cerebral pathology. Brain involvement in FD is thought to be mainly mediated by lysosomal deposition of catabolites in endothelial cells, leading to micro- and macro-vascular manifestations that partly overlap with those occurring in common small vessel disease and normal aging.(23) Uncertainty exists on the relative importance of a possible direct brain tissue damage through lysosomal deposition at the level of other cell types (i.e., neuronal or glial cells).(24) Interestingly, the accumulation of lysosomal storage bodies in microglia subsets is a physiological process that increases linearly with aging, suggesting a convergence of mechanisms operating during normal aging and in lysosomal storage disorders, with the accumulation of storage bodies potentially driving microglia dysfunction and contributing to neurodegeneration.(25) Taken together, these observations reinforce the analogy between FD and accelerated aging, with brain-PAD as a plausible neuroimaging biomarker of progressive FD-related brain damage. On the other hand, FD has been also described as a neurodevelopmental disorder,(11) and brain-age predictions are known to be influenced by early-life factors related to brain development.(26) Therefore, we cannot exclude that abnormal neurodevelopment might play a role in the observed group differences in terms of brain-predicted age.

The clinical relevance of brain-PAD in FD is illustrated by a significant association with overall, multi-organ, clinical severity in patients with FD. An intricate network of mutual interdependencies exists between brain-age and other bodily (patho)physiological aging phenomena.(27,28) Brain health is shaped by other systems in the body, with cardiovascular and renal (mal)functioning known to have a potentially major impact.(7,29,30) Likewise, common, genetically-determined, mechanisms may underlie the development of FD-related damage in different organs in parallel.

From a neuroimaging perspective, brain-age predictions in patients with FD were influenced by WMH and brain volume, with no anatomical specificity other than greater effect sizes observed at the level of the deep and periventricular WM, which are known to be preferentially impacted by FD-related lesions and microstructural damage.(31) Our results are in line with previous evidence, confirming the role of a smaller brain volume, rather than the atrophy of specific regions, and a greater WMH burden as the main neuroimaging correlates of higher brain-PAD values,(7,21,22) with no FD-specific determinants emerging from the interaction analysis. Interestingly, several structural MRI studies using more traditional approaches (i.e., voxel-based morphometry and region of interest-based analyses) have failed to reveal consistent changes of brain morphometry in FD patients compared to HC,(3,10,32) at least partially because of the small sample sizes. In this light, brain-PAD might represent a more comprehensive biomarker of brain structural health in FD, potentially more sensitive than conventional methods.

Overall, our findings support the role of brain-PAD as a sensitive quantitative biomarker of FD severity, with potentially relevant implications for patient stratification in clinical and research settings, especially in relation to the assessment of treatment response. Indeed, the efficacy of recently introduced specific treatments for FD on cerebral involvement has remained unclear so far, partly because of the very lack of objective neuroimaging measures of disease severity.(33,34)

The main limitation of our study lies in its cross-sectional nature. As mentioned, disentangling the relative effects of neurodevelopmental factors and ongoing pathological processes on brain-PAD is challenging in a cross-sectional setting. Assuming that the impact of neurodevelopment remains constant over time, longitudinal studies are warranted, where brain-predicetd age deltas ideally depend solely on ongoing healthy/pathological aging phenomena. Also, a longitudinal design would allow to assess the prognostic value of brain-PAD towards clinical outcomes. Furthermore, crossing brain MRI-derived metrics with additional, more specific, biomarkers of disease severity (e.g., plasma Lyso-Gb3 levels)(35) would help to link brain-PAD more closely to FD pathogenesis.

In conclusion, we provide evidence that the brains of patients with FD appear older than normal. The brain-PAD metric is sensitive to FD-related brain damage and is associated with systemic involvement, positioning it as a relevant quantitative imaging biomarker to assess (neurological) disease severity in clinical and research settings.

## Data Availability

The data that support the findings of this study are available from the corresponding author upon reasonable request.

## Supplemental Materials

**Supplementary Table 1.** Data sources for healthy brain age model training and validation.

| Cohort | N | Age mean (SD) | Age range | Sex male/female | Repository details | Scanner (Field strength) | Scan | Voxel dimensions |
| --- | --- | --- | --- | --- | --- | --- | --- | --- |
| **ABIDE (Autism Brain Imaging Data Exchange)** | 557 | 16.90 (7.62) | 6-56 | 461/96 | INDI | Various (all 3T) | MPRAGE | Various |
| **Berlin School of Brain & Mind** | 49 | 31 (7.11) | 20-60 | 24/25 | INDI | Siemens Tim Trio (3T) | MPRAGE | 1.0 x 1.0 x 1.0 |
| **Cleveland Clinic** | 31 | 43.55 (11.13) | 24-60 | 11/20 | INDI | Siemens Tim Trio (3T) | MPRAGE | 2.0 x 1.0 x 1.2 |
| **International Consortium for Brain Mapping (ICBM)** | 640 | 30.63 (12.17) | 18-80 | 349/291 | LONI IDA | Siemens Magnetom (1.5T) | MPRAGE | 1.0 x 1.0 x 1.0 |
| **Information eXtraction from Images (IXI)** | 564 | 48.66 (16.53) | 20-86 | 252/312 | http://brain-development.org/ixi-dataset/ | Philips Intera (3T); Philips Gyroscan Intera (1.5T); GE Signa (1.5T) | T1-FFE; MPRAGE | 0.9375 x 0.9375 x 1.2 |
| **MIND Clinical Imaging Consortium (MCIC)** | 95 | 32.89 (12.17) | 18-60 | 65/30 | COINS | Siemens Sonata/Trio (1.5/3T); GE Signa (1.5T) | MPRAGE; SPGR | 0.625 x 0.625 x 1.5 |
| **Minimal Interval Resonance Imaging in Alzheimer's Disease (MIRIAD)** | 23 | 69.66 (7.18) | 58-85 | 12/11 | https://www.ucl.ac.uk/drc/research/methods/minimal-interval-resonance-imaging-alzheimers-disease-miriad | GE Signa (1.5T) | 3D IR-FSPGR | 0.9375 x 0.93751 x1.5 |
| **Nathan Kline Institute (NKI) / Rockland sample** | 201 | 35.30 (20.01) | 4-85 | 119/82 | INDI | Siemens Tim Trio (3T) | MPRAGE | 1.0 x 1.0 x 1.0 |
| **Brain-age modelling cohort** | 2160 | 32.94 (18.06) | 4-86 | 1293/867 | - | - | - | - |

INDI = International Neuroimaging Data-sharing Initiative (http://fcon_1000.projects.nitrc.org)
COINS = Collaborative Informatics and Neuroimaging Suite (http://coins.mrn.org)
LONI = Laboratory of Neuro Imaging Image & Data Archive (https://ida.loni.usc.edu/)
ABIDE consortiums comprising data from various sites with different scanners/parameters

**Supplementary Figure 1.**
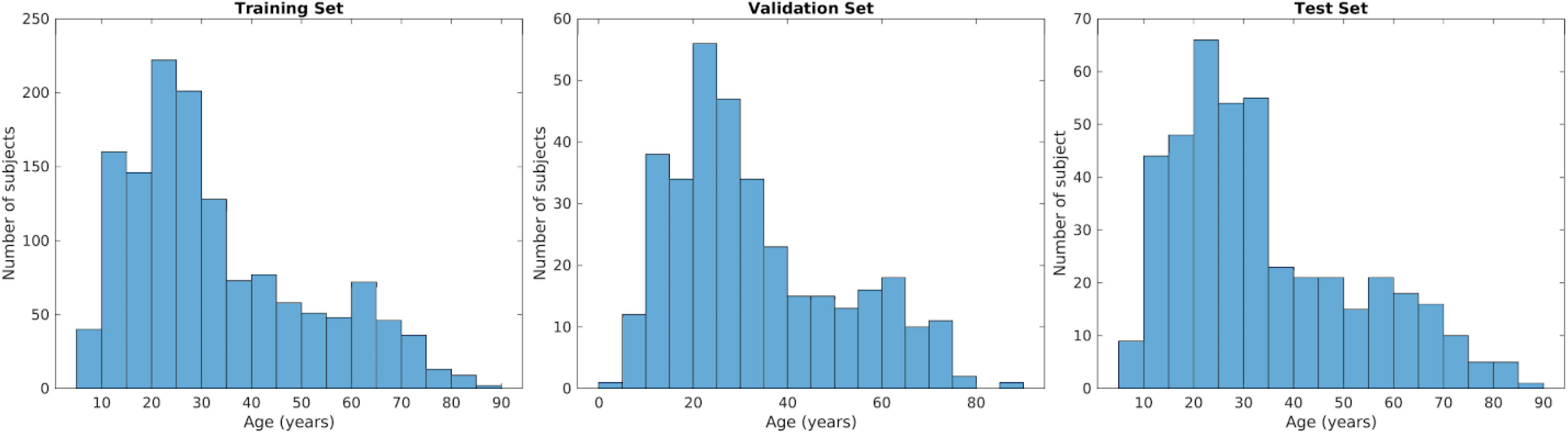
Age distribution in training, validation, and test sets.

